# Semi-automated socio-anthropologic analysis of the *medical discourse* on rheumatoid arthritis: potential impact in public health

**DOI:** 10.1101/2022.04.05.22273443

**Authors:** Christine Nardini, Lucia Candelise, Mauro Turrini, Olga Addimanda

## Abstract

The debilitating effects of non-communicable diseases (NCDs) and the accompanying chronic inflammation, represent a significant obstacle for the sustainability of our development, with efforts being spread worldwide to contrast NCDs’ diffusion, as per the United Nations Sustainable Development Goals (SDG 3). In fact, despite efforts of variable intensities in numerous directions (from innovations in biotechnology to lifestyle modifications), NCDs’ incidence remains pandemic.

The present work wants to contribute to this major concern with a specific focus on the fragmentation of the medical approaches, via an interdisciplinary analysis of the *medical discourse*, i.e. the heterogenous reporting that biomedical scientific literature uses to describe the anti-inflammatory therapeutic landscape in NCDs. The aim is to better capture the roots of this compartmentalization and the power relations existing among three segregated *pharmacological, experimental* and *unstandardized* biomedical approaches, to ultimately empower collaboration beyond medical specialties and possibly untap a more ample and effective reservoir of integrated therapeutic opportunities.

Using as exemplar disease rheumatoid arthritis (RA), twenty-eight articles were manually translated each into a nine-dimensional categorical variable of medical socio-anthropological relevance, relating in particular (but not only) to *legitimacy, temporality* and *spatialization*. This digitalized picture (9 × 28 table) of the medical discourse was further analyzed by simple automatic learning approaches to identify differences and highlight commonalities among the biomedical categories.

Interpretation of these results gives original insights including the suggestion to: empower scientific communication between *unstandardized* approaches and basic biology; promote non-pharmacological therapies repurposing to enhance robustness of *experimental* approaches; align the spatial representation of diseases and therapies in *pharmacology* to effectively embrace the *systemic* approach promoted by modern personalized and preventive medicines. We hope this original work may expand and foster interdisciplinarity among public health stakeholders, ultimately contributing to the achievement of SGD3.

## Introduction

Chronic inflammation is a known trigger and symptom accompanying non-communicable diseases (NCDs). NCDs are responsible for 40 million death per year and have been recognized a silent pandemic [1] long before the dramatic rise of covid-19 transmissible infection. Chronic inflammation starts as a subtle alteration of the inflammatory response, whose impact remains covered for years before the effects become overt and allow a clear diagnosis of the accompanying malady. Therefore, the management of chronic inflammation represents an important component as well as a tremendous burden for sustainable development, as clarified by the ambition of the United Nations Sustainable Development Goals (namely, SDG 3, [2]) and further regional [3] and national implementations [4].

In this frame, several approaches are being defined and explored to leverage on the problem from biomedical research to identify novel and more effective anti-inflammatory therapies [5], to health policy strategies to cope with low-cost high-impact solutions [6]. Our aim with this study is to contribute from yet another viewpoint, i.e. by analyzing how NCDs’ management is currently conceived and presented, using as proxy to gain insight into this issue an interdisciplinary analysis of the *medical discourse*. The *medical discourse* defines how medical practitioners and researchers (or better, a core group of experts committed to research on clinical practice) appraise scientifically the best therapeutic options available for a specific condition [7]. Contrary and somewhat surprisingly to the biomedical understanding this approach is far from uniform or standardized. Indeed, the medical discourse provides an arena open enough to host the plurality of conceptions of a disease as well as the techniques and practices to treat it, along with the power relations among these different approaches, ultimately defining which therapies will be offered in the clinical routine.

Studies at the intersection of anthropology, sociology of science and medicine, have shown how such (pre)conceptions shape both biomedical research and health policy strategies for example in the etiological account of a genetic disease [8], the regulation of a genetic test [9], or in the usage of support tools for medical decision-making [10]. In the current work, analyses of socio-anthropological categories are devised to explore the dichotomous labeling that characterizes the current therapeutic offer (*pharmacological* vs *non-pharmacological, mainstream* vs *experimental, standardized vs unstandardized*). Given the complexity of the task and its vastness, we narrow our focus along three lines.

First, our analysis is run from the *biomedical* standpoint, i.e. we explore the medical discourse as it is presented in *biomedicine*. Biomedicine, despite its numerous flavors [11], is generally understood as “the branch of medicine concerned with the application of the principles of biology and biochemistry to medical research or practice” [12], including by now bioinformatics, computational and systems biology [13] as well as network medicine [14,15], and we use this term to indicate the most extensively recognized medical approach, in other words, the *clinical gold standard*. To take this into account we use Pubmed (https://pubmed.ncbi.nlm.nih.gov/) as our reference data base. Second, as an exemplar malady to drive our analysis we use rheumatoid arthritis (RA) a recognized model pathology for chronic, inflammatory autoimmune diseases, keeping in mind that while this choice makes the discussion more manageable, all related considerations are, in principle, easily translatable to other NCDs. In practice, this implies that the therapeutic gold standard is defined by regional associations like the American College of Rheumatology (ACR [16]), the EUropean and Asian Pacific Leagues Against Rheumatisms EULAR [17] and APLAR [18]. And include drugs that aim at the symptomatic control of inflammation like *non-steroideal anti-inflammatory* drugs (NSAIDS), and paracetamol or morphine(-derived) *analgesic* drugs; *corticosteroid* which operate beyond symptoms and attempt to contrast the degeneration associated with the disease; and finally *disease modifying anti-rheumatic drugs* (DMARDs) able to interfere and block on more (biologic, bDMARDs) or less (conventional, cDMARDs) specific targets, the activity of pro-inflammatory molecules (cytokines) or cells, once the disease has been diagnosed [16–18]. Third, to identify RA available therapeutic options, beyond the biomedical pharmacological gold standard (from now on *PHA*), we use the framework represented by the *greater inflammatory pathway* (GIP,[19]) a recent conceptualization of inflammation which includes in addition to the immune response: (i) the role of the gut-intestinal (GI) microbiome, (ii) the role of the autonomous nervous system (ANS); (iii) the reactions to physical stimuli described under the umbrella of *wound healing*. This represents a blueprint to build our queries on Pubmed including all RA therapies modulating the GIP functions, which therefore account for: (i) modulators of the *GI microbiome*: diet, nutraceutics, antibiotics and fecal microbial transplant (FMT); (ii) modulators of the *ANS*, which coincides to date with vagus nerve stimulation (VNS), which jointly represent novel experimental therapies (*EXP*) and (iii) *wound healing* modulators in the form of optical, mechanical, magnetic and electrical stimulation-based therapeutic approaches, offering limited standardization, evidence-based research, guidelines, funding and overall public recognition, despite widespread use. We name these latter approaches *unstandardized non-pharmacological (USTD)*.

The source we use to appraise our analysis (see the overview Fig 1) is represented by scientific reviews, a privileged source for the qualitative analysis of biomedical practices or techniques. Reviews, based on a selection of existing research papers, are committed to elaborate a factual evaluation of a dedicated topic. Accordingly, they tend to hide differences and ambiguity and define, more or less implicitly, the legitimate boundaries of a specific practice or technique [24,25]. Our effort is specifically to make these aspects explicit, through the lenses of a number (*nine*) of socio-anthropological categories. Namely, we extracted from each review the *typology* of the article (we distinguished *systematic reviews* i.e. data driven statistical elaboration of multiple clinical studies, and *overviews* that collect in a less systematic way different types of scientific publications, including pilot and exploratory studies); the *temporality* at stake [26] (meaning whether the treatments have a long dated practice in the biomedical environment -*experiential*-, or whether they are *innovations* based on a promissory areas of investigation); their *legitimacy* vis-à-vis the dominant pharmacological paradigm [27,28] (meaning *alternative* or *complementary* in relation with other approaches) and the *spatializations* in the body [26,29] of the disease’s *etiology* and *treatment* (*local* or *systemic*), i.e. where, in the body, the malady or the therapy are thought to be operating. In addition we collected the diagnostic *criteria* as well as disease activity indices or health assessment parameters (ACR/EULAR [30], OMERACT[31] or Range of Motion, ROM, i.e. the measurement of movement around a specific joint), the *authors’ background* (extracted by the proxy of the main author’s *affiliation*); the *year* of publication and the *geographic* localization (extracted from the nation where the institute of affiliation is located, as a loose proxy of cultural environment). Full details are given in Methods and Table 1.

**Table 1.**
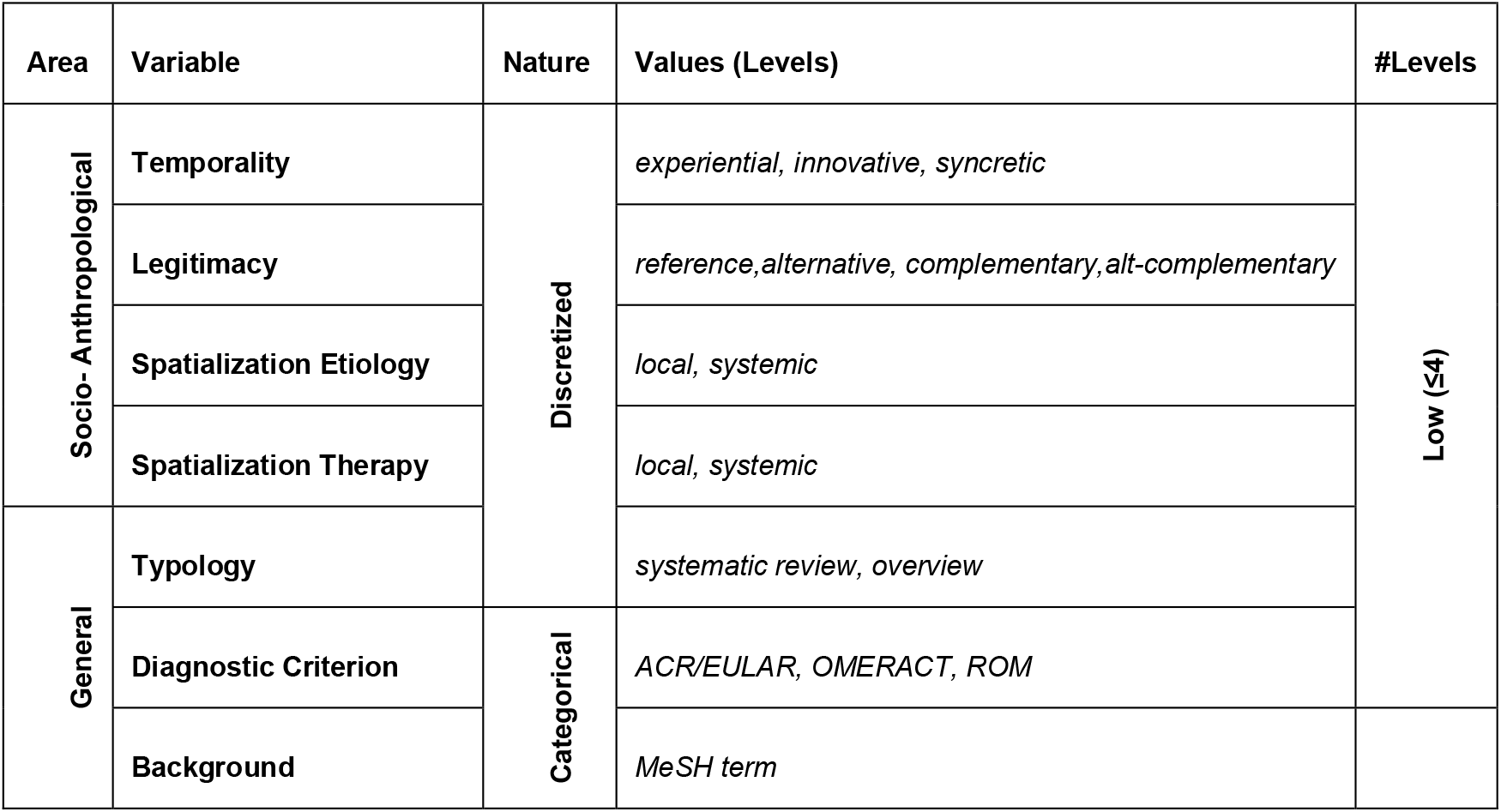
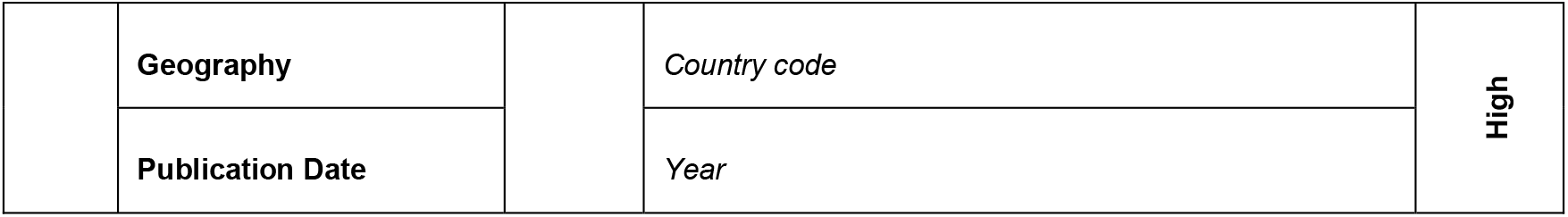
Variables characterizing the study.

**Figure 1.**
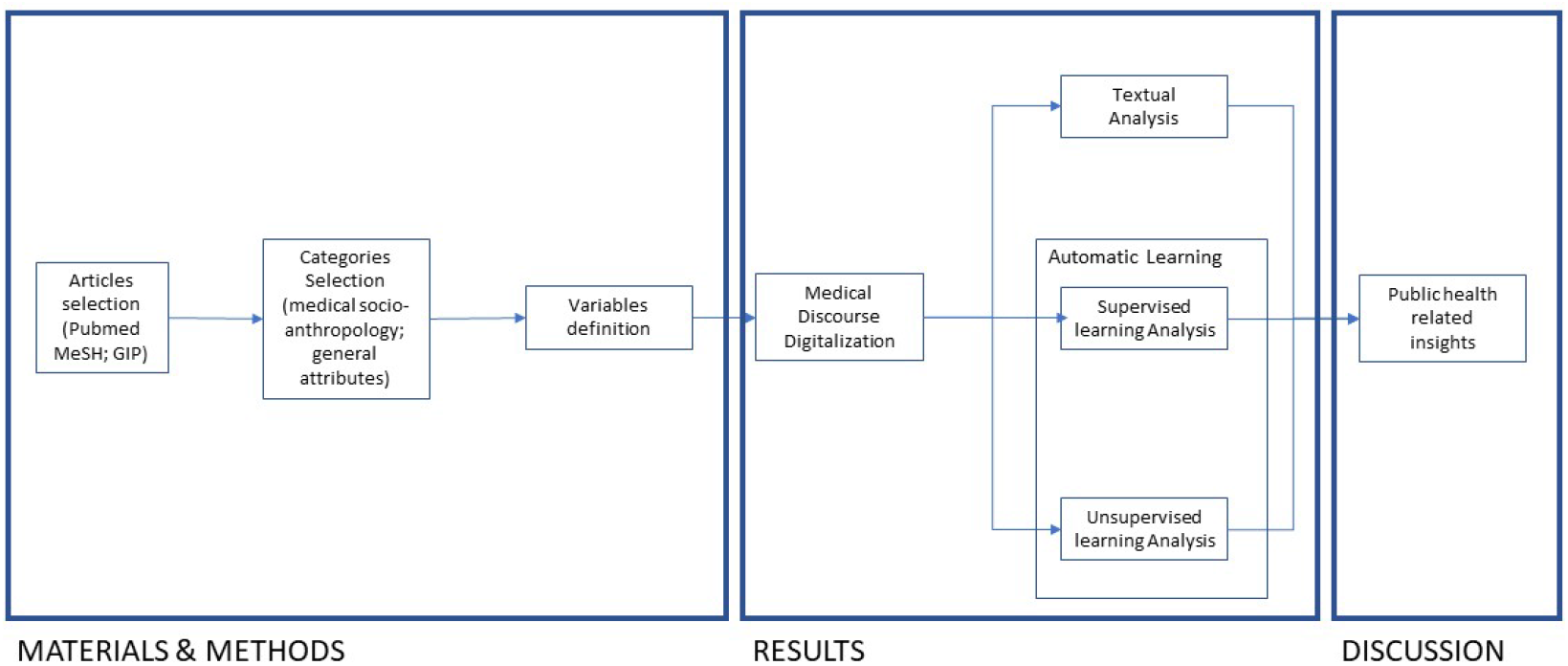
Schematic representation of the phases of the analysis of the medical discourse on RA and the corresponding sections of this article.

Finally, manual curation of all such categorical variables, article-wise, is merged into a tabular format (Table 2) representing a global, digitalized picture of the medical discourse. This type of representation enables, in addition to the more common medical socio-anthropological *textual* analysis, the usage of automatic tools for investigation, and in particular the so called *supervised* and *unsupervised* machine learning approaches. The former (*supervised*) uses *a priori* knowledge on the data, this implies that the analysis is run in search of a (novel) representation of a category, variable, cluster or concept of interest, known *a priori*. In our case, this is represented for each of the articles of our collection by its *biomedical category (PHA, EXPT, USTD)*, with the aim to give each category an original socio-anthropological interpretation. The latter (*unsupervised*) allows a *systemic* view on the medical discourse on RA, letting appear patterns (or *emergent properties*) that would hardly be visible by an analysis run article-, category- or therapy-wise, and thus offering what we hope to be an original, insightful critique on the state-of-the-art, opening to original and fruitful considerations for public health stakeholders.

**Table 2.** Digitalized summary of the medical discourse

| Cat. | Label | Authors | Typology | Temporality | Sp. Etiology | Sp. Therapy | Legitimacy | Criterion | Date | Background (MeSH) | Geo |
| --- | --- | --- | --- | --- | --- | --- | --- | --- | --- | --- | --- |
| PHA | AI | Burmester GR et al. | sys. rev. | innovation | localized | localized | reference | ACR/EULAR | 2017 | Rheumatology | DE |
| PHA | AI | Ramiro S et al. | sys. rev. | innovation | localized | localized | reference | ACR/EULAR | 2012 | Rheumatology | PT |
| PHA | AI | Gaffo A et al. | sys. rev. | innovation | localized | localized | reference | ACR/EULAR | 2006 | Rheumatology | US |
| PHA | AI | Konttinen YT et al. | sys. rev. | innovation | localized | localized | reference | ACR/EULAR | 2005 | Medicine | FI |
| PHA | AI | Lee SJ et al. | sys. rev. | innovation | localized | systemic | reference | ACR/EULAR | 2003 | Rheumatology | US |
| PHA | AI | Fries JF | sys. rev. | innovation | localized | systemic | reference | NA | 2000 | Rheumatology | US |
| PHA | AI | Porter DR et al. | overview | experiential | localized | systemic | reference | NA | 1993 | Rheumatology | UK |
| PHA | AI | Huskisson EC et al. | overview | experiential | localized | systemic | reference | NA | 1978 | Rheumatology | UK |
| EXP | VNS | Koopman FA et al. | overview | innovation | systemic | localized | reference | ACR/EULAR | 2017 | Rheumatology | NL |
| EXP | VNS | Koopman FA et al. | overview | innovation | systemic | localized | reference | ACR/EULAR | 2014 | Rheumatology | NL |
| EXP | Dys | Jethwa H et al. | overview | innovation | systemic | systemic | complementary | NA | 2017 | Medicine | UK |
| EXP | Dys | Kang Y et al. | overview | innovation | systemic | systemic | complementary | NA | 2017 | Inf. Dis. Medicine | CN |
| EXP | AB | Mottram PL | overview | experiential | localized | localized | complementary | NA | 2003 | Medicine | AU |
| EXP | AB | Kloppenburger et al. | overview | experiential | localized | localized | complementary | NA | 1995 | Rheumatology | NL |
| EXP | Diet | Schorpion A et al. | overview | innovation | systemic | systemic | complementary | ACR/EULAR | 2017 | Rheumatology | US |
| EXP | Diet | Rennie KL et al. | overview | innovation | systemic | systemic | complementary | NA | 2003 | Nutritional Sciences | UK |
| EXP | FMT | Zhang X. et al. | overview | innovation | systemic | localized | complementary | ACR/EULAR | 2019 | Rheumatology | CN |
| USTD | US | Beasley J. | sys. rev. | experiential | localized | localized | complementary | NA | 2012 | Occupational Med. | US |
| USTD | US | Casimiro L et al. | sys. rev. | experiential | localized | localized | complementary | ACR/OMERACT | 2002 | Phys.& Rehab. Med. | CA |
| USTD | Mass | Nelson NL et al. | sys. rev. | experiential | systemic | systemic | alternative | WOMAC/OMERACT | 2017 | Applied Kinesiology | US |
| USTD | AP | Lee MS et al. | overview | experiential | systemic | systemic | alt-complementary | ACR/EULAR | 2008 | Oriental Medicine | KR |
| USTD | AP | Casimiro L et al. | sys. rev. | experiential | systemic | systemic | complementary | WOMAC/OMERACT | 2005 | Phys.& Rehab. Med. | CA |
| USTD | AP | Wang C et al. | overview | experiential | systemic | systemic | alt-complementary | ACR/EULAR | 2008 | Rheumatology | US |
| USTD | EL | Brosseau L et al. | sys. rev. | experiential | localized | localized | complementary | WOMAC/OMERACT | 2003 | Phys.& Rehab. Med. | CA |
| USTD | EL | Pelland L et al. | sys. rev. | experiential | localized | localized | alternative | NA | 2002 | Physiotherapy | CA |
| USTD | LLT | Brosseau L et al. | sys. rev. | experiential | localized | localized | complementary | WOMAC/OMERACT | 2005 | Phys.& Rehab. Med. | CA |
| USTD | LLT | Christie A et al. | sys. rev. | experiential | systemic | localized | complementary | WOMAC/OMERACT | 2007 | Rheumatology | NO |
| USTD | EM | Ganesan K et al. | overview | syncretic | systemic | systemic | alternative | NA | 2009 | Biotechnology | IN |

Clearly our analysis suffers from the overall limited number of articles we used, resulting from the necessary manual curation to identify the variables of interest (expansion and criticism towards higher automation of this analysis may be the object of future work [32]). Further, our attention is turned here to the perception of *non-pharmacological* therapies, for their potential cost-effectiveness (with an eye at *frugal innovation* [33] given the numbers and costs at stake), for this reasons other *pharmacological experimental* therapies are not taken into account at this stage (for instance, but not limited to, delivery by nanoparticles or stem cells [5]).

Nevertheless, despite the room that this analysis leaves to broader and more complete explorations (including the promise of a soon-to-come ACR document on the non-pharmacological approaches to RA [16]), we consider the literature extracted by our criteria as being part of the interest of this study. In fact, the amount and type of articles on a given topic are also function of its acceptance and recognition by the community, including perceived interest, relevance and funding opportunities, to name a few.

## Materials and methods

### Materials - reviews

Twenty-eight reviews were retained upon a literature search of PubMed run using Medical Subject Headings (MeSH [34]) to compose a three-parts query. The first part, common to all queries broadly searches for “Arthritis, Rheumatoid/therapy”[Mesh]) AND “Review” [Publication Type]”. The second and third queries refine the first with specifiers including: (level I) the modulators of the GIP [19], and their physical nature (level II), see Fig 2 for dependencies and S1 Table for the complete text of the queries. Level I includes as summarized in the introduction: gut intestinal microbiome, autonomous nervous system, wound healing. Level II is taken by the list of stimuli (optical, mechanical, electric, magnetic) that are routinely used in wound healing testing [35], casting heat within mechanical stimulation. The list of results obtained by all queries was narrowed down by excluding reviews that were: not on RA; not in English; not appropriate; too specific; too general, or misclassified (see Supporting Information).

**Figure 2.**
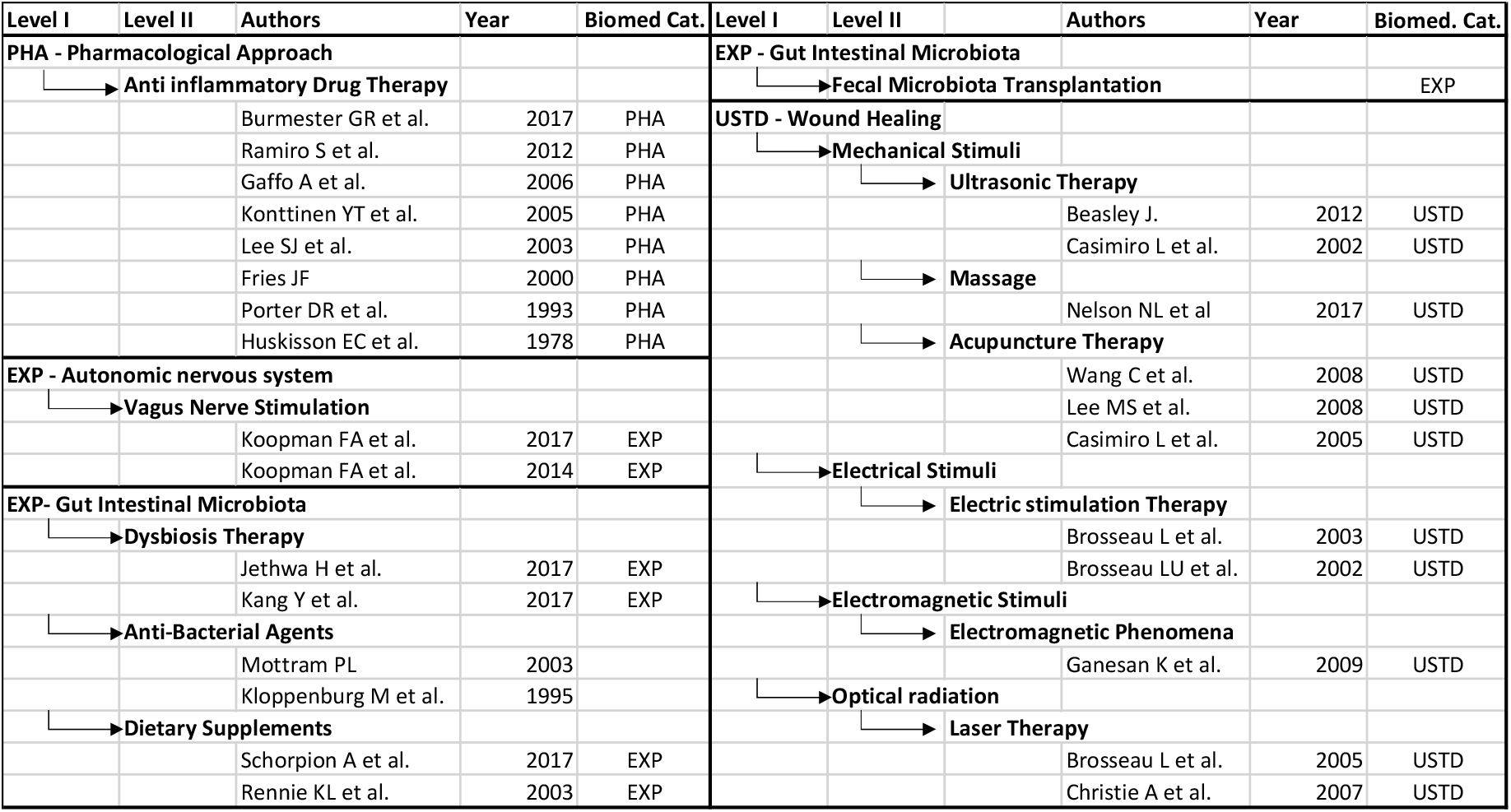
List of publications (first author, date of publication) grouped by query. Owing to space constraints only minimal information have been retained, full data are collected for inspection in Supplementary Materials and S1-2 Tables directly importing MeSH database PubMed search. * The article does not have a doi, PMIC is used instead. * *no article was found in relation with FMT and RA, however a clinical trial is ongoing.

The number of articles resulting from each query varied widely, ranging from three items for VNS (electrical stimulation of the ANS) to hundreds for pharmacological anti-inflammatory drugs. The list was finally manually curated to retain the two most recent and representative reviews for each therapeutic approach. There are some exceptions (see details in Supporting Information): (i) for the query on *anti-inflammatory drugs*, which represents the dominant biomedical approach to RA and offers, alone, a sufficiently long temporal excursus, i.e. an historical perspective on its evolution, we preserved 8 articles; (ii) for acupuncture in addition to the Cochrane review [36] which we preferentially selected when available, we preserved two more coeval articles [37,38], building on virtually the same material (meta-analysis of 8 clinical study, 6 of which in common, and both including the only 2 studies preserved in the Cochrane review) yet characterized by a very different *discourse* and medical conclusions; (iii) for magnetism and massage only one article could be retrieved with the above defined criteria; (iv) no article on FMT applied to RA was available, however a clinical study is ongoing (NCT03944096). Identification of the variables levels (see below) was inferred where not available in the trial record from other FMT studies [23]. For the sake of readability, from now on all 28 items of our selection are referred to as *articles*. Detailed results are included in S2 Table.

### Materials - Variables

The identification of the variables relevant to handle the analysis followed several steps. First, upon careful study of the available literature, it was considered appropriate to identify categories from the areas of medical sociology [39] and anthropology [40], as well as some general attributes, to capture the essence of the *medical discourse* expressed in the selected material. Table 1 reports this step in column *Area*. Second, intense discussion identified the categories that best captured in socio-anthropological terms the observed ample variance among the articles, namely the *Temporality* [26], *Legitimacy* [27,28] and *Spatialization* of the approaches [26,29], backed by the additional attributes: *Typology* of the article, Diagnostic *Criterion, Background* of the authors, *Date* of Publication, *Geography* (column *Variables*). Third, in order to go beyond the study of individual articles and make the process of analysis more manageable, variables that were not inherently categorical, where discretized (column referring to the mathematical *Nature* of the variable). This additional step implied a standardization of the values of each variable (column *Values*). Values were assessed by a sociologist and an anthropologist for the socio-anthropological variables each on a subset of articles, and by a systems biologist for the general variables and for inference on the FMT study, using the controlled MeSH vocabulary [34], where appropriate. Finally, variables were observed for the dimensionality of their level (column *#Levels*), with variables characterized by a number of levels comparable to the sample size (twenty-eight) being further processed by *textual analysis*, and the other by *automatic learning* (see Methods).

In the following: *variables* represents also the *dimensions* of the socio-anthropological space, and *values* of the categorical variables are interchangeably referred to as *levels*. All articles were read and final values were discussed and agreed upon by all researchers.

*List of the 9 categorical variables used in this article. From left to right, the first column identifies the* Area *in which the categories useful to describe the articles’ medical discourse were selected. Column* Variable *identifies the names taken or derived from the corresponding socio-anthropological categories or general attributes; the third column reports the mathematical* Nature *of the qualitative variables so defined; column* Values *reports the result of the process of discretization, or the levels of the categorical variables; column* #Levels *points to the dimensionality of the variable values, a criterion that guides the analysis presented in Results (textual or automated). Follow the definitions of the nine selected variables*.

1. *Temporality*. The two main sources of scientific soundness – experiential knowledge and technoscientific innovation *–* are inscribed within two opposing temporal polarities: past and future [26]. *Experiential* knowledge is associated with established practices, that have been adopted and proven effective within comparable clinical frames for a sufficiently long time in the (recent) past. Technoscientific *innovation* is associated to new tendencies within biomedical research. In such cases, a treatment is deemed promising in light of the results it has produced in basic research, *in vitro, in vivo* or in early stages of clinical validation. An exception, the very peculiar and unique article on electromagnetism [41], forced us to add an intermediate level: *syncretism*.
2. *Legitimacy*: describes the relationship of a therapy considered vis-à-vis the dominant pharmacological approach [27]. This category is intended to capture whether a therapeutic approach proposes itself as an alternative or complementary remedy. We chose to use these two categories that refer to the now popular definition given in the 1990s by the US National Institutes of Health [42], where the definition of Complementary and Alternative Medicine (CAM). In our work we use the terms *complementary* or *alternative* to indicate the *relative* relationship that a given treatment has with any other treatment, not to biomedicine alone. Generally, we speak of complementary or alternative *with respect to* biomedicine. However, in some cases this relationship refers to different treatments [43]. In few cases [37,38] it was necessary to introduce an additional intermediate value for the dual/ambiguous relationship with other medical approaches, using *alt-complementary*. The literature on anti-inflammatory drugs represents the *reference* and is labeled as such.
3. S*patialization of the disease’s etiology*. This variable analyzes in which region of the body the medical discourse locates the disease’s origin (etiology) [29]. Modern medicine generally makes the disease evolve from an original place, the *lesion*, located at a very precise point in the depth of the organs and verifiable through autopsy [44]. Some authors [45] advanced the idea that in contemporary biomedicine the *lesion* is represented at the molecular level, as the interaction between enzymes, proteins and other biochemical or cellular elements. This is the case of the dominant PHA approach to RA, according to which this condition descends from a molecular, immunological abnormality (partly genetic and partly environmental) that leads to an autoimmune response, inflammation, senescence, and, finally, to the joints damage. Other approaches, however, adopt a more holistic vision of the body and, accordingly, interpret pathology as a more general condition, involving interactions between different organs, or relationships with the outside (for instance food). Traditional approaches usually tend to be holistic, yet, within the biomedical world, systemic approaches to the body can also be found, as it is the case for the role of the GI microbiome. With regard to body geographies, the distinction was made between *systemic* approaches, which conceive the disease as a widespread phenomenon in the organism and arising from the interaction of several organs and functional systems, or *localized*, in the case of approaches that understand the disease as the result of a lesion located in a specific area.
4. *Spatialization of the disease’s therapy*. In our pool of articles we observed that the same rationale described above could be applied to the spatialization of the therapy, as the two (spatialization of the etiology *or* of the disease) did not necessarily overlap. A therapy is systemic when it considers the healing processes as involving the whole organism, or the interaction with the environment (as it is the case for nutrition), or different regions of the body (as it is the case for VNS or dysbiosis), or holistic body descriptors (such as *meridians* in Traditional Chinese Medicine). When the perception of the disease and of the therapy spatially coincide (i.e. the values of the two spatialization variables overlap), meaning if they are both *local* or both *systemic*, we define them as *coherent* and *incoherent* otherwise.
5. *Typology:* This criterion has emerged with force from the manual curation of the articles. Despite all articles belonging to the biomedical scientific literature (all responding to the definition of ‘review’ as per the MeSH search term) they consider a wide set of different articles and adopt different analytical approaches. In particular, we chose to make a distinction between *systematic review* and *overview. Systematic reviews* are committed to analyze the impact of a specific treatment based on human clinical trials selected through specific criteria and they include (but are not limited to) statistical *meta-analyses*, whereas *overviews* collect in a less systematic way different types of scientific publications (clinical trials, but also *in vitro* or animal studies) on the basis of the authors’ knowledge of the state-of-the-art with the aim to offer insights about a particular technique or research branch.
6. Diagnostic *Criterion*. This refers to the description of the diagnosis and disease progression, against a list of standardized parameters. These are the definition regularly updated by the American Congress of Rheumatology (ACR) and European League for Arthritis Rheumatoid, EULAR, often also referred jointly as ACR/EULAR [30], relying on clinical, molecular as well as functional parameters assessed by patients and doctors (using in particular the Disease Activity Score, DAS28 to objectively assess the number of affected joints [46]), and the OMERACT ([31] consensus-derived collection of outcomes and instruments to measure a consistent set of clinical endpoints in RA, with a particular focus on the damage assessed by ultrasound). Finally, ROM (range of motion score, to measure the distance and direction that a joint can stretch) was used exclusively in the review on massage [47] In one case only two indexes are used jointly (ACR OMERACT, ultrasound therapy [48])
7. *Background* of the authors, using as proxy the MeSH term corresponding to the specialty of the Department or (where absent) of the Institute of affiliation.
8. *Date of Publication*, by year.
9. *Geography* using the State of the main author’s institution of affiliation. This has been used as a loose proxy for the cultural context the article emerges from.

## Methods Analysis

Once each of the twenty-eight articles has been transformed in a vector 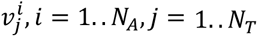, with *N*_*A*_ = 28 number of articles selected, and *N*_*T*_ = 9 total number of variables, juxtaposition of such vectors offers the digitalized representation of the medical discourse shown in Table 2, further explored via two main approaches: textual analysis and automatic learning.

### Textual Analysis

For categorical variables characterized by a number of levels comparable to the size of the dataset (variables share almost the same dimensionality of the space, for Background, Date, Geography) automatic learning is meaningless and hence textual socio-anthropological analysis was preferred. This represents an highlight and a dedicated deepening of the extensive work of manual curation that underlie the discretization of *all* articles for *all* variables we retained. This approach was also deemed useful as a contextualized introduction for the following automatic learning analyses.

### Automatic learning

For the six variables characterized by a couple to a handful of levels (Typology, Temporality, Spatialization Etiology and Therapy, Criterion and Legitimacy), automatic analysis was possible, and run with two further subsets of approaches.

First, to identify for each of the three *biomedical categories* (*PHA, EXP, USTD*) a *representative* vector, we used as supervised approach the simplest form of *majority voting*.

In brief, for each biomedical category *C*_*i*_, *i* = 1.. *N*_*C*_, *N*_*c*_ = 3, each retained socio-anthropological variable *v*_*j*_, *j* = 1.. *N*_*v*_, *N*_*v*_ = 6 and each level (value) of the categorical variable 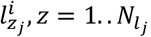, with 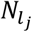 the number of levels in variable *v*_*j*_, the *N*_c_ = 3 *reference* vectors’ values 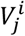 are built as follows:

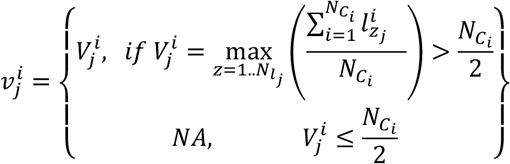

i.e., in the reference vector, the variable’s level is chosen as the one that has the majority frequency (if any) across all articles in the given category. When the absolute majority (frequency above half of the cases) does not exist the value is left empty (NA).

Second, to search for *emergent properties*, given the low dimensionality of the space and the dataset, patterns were searched by visual inspection of the clusters identified by the variables’ graphical distributions on a scatter plot, mapping the six variables of interest to the x and y axes (Typology and Temporality), using facets to break the two Spatializations (Etiology and Therapy) and having points’ shapes and colors to map Criterion and Legitimacy, respectively. All plots and analyses were run in R using packages *ggplot2* [49] and *superheat* [50].

## Results

The methodology described above offers as first result the manually curated translation of each of the twenty-eight articles into a nine-dimensional categorical variables vector, i.e. the digitalized picture of the medical discourse on the anti-inflammatory treatments on RA shown in Table 2. Its analysis proceeds in three steps shown in Fig 1.

First, an individual variables’ discussion is offered (textual analysis), with respect of the *Date* of publication, *Background* of the authors and *Geography* (see Methods). Second, the automated processing (via simple machine learning) of the six other variables as a whole (system) allows to identify different types of *emergent* properties defining the biomedical categories from a novel socio-anthropological perspective and to gain original insight in their power relations (via supervised and unsupervised analysis).

## Textual analysis - individual variables

### Background

Authors’ background play different roles in the presentation of treatments. At one end of the spectrum there are authors pivotal in actively supporting (innovative) therapies as it is the case for VNS (whose authors are pioneers in the approach, hence *overviews* mostly) or nutrition (where nutritionists explain how this medical specialty should be interested in patients who already, in an autonomous and amateurish way, are interested in taking a suitable diet). At the opposite end (generally for therapies that are *experiential*, as well as for pharmacological treatments), we have *systematic reviews*, in most cases edited by the Cochrane Musculoskeletal group around the 2000s (between 2002 and 2005) and written by the same authors (three authors appear in all five articles we selected). It is particularly interesting to note that among these authors one only appears to be working in a rheumatology department, and that none of the other authors presents an affiliation that is directly involved in the treatments examined. In these cases, therefore, the *systematic overview* collects and compare statistical data from clinical studies evaluated on the basis of formal criteria and without any practical knowledge of the dynamics associated with the treatment, except mainstream therapies.

### Geography

The geographic origin and the knowledge of a language also affect the selection of the data used to build the articles. Namely, the article on drug therapies published in 2005 [51] by a group of researchers from Finland often points out that the presented approach is specific to northern European countries; the Cochrane Review on acupuncture [36] includes only articles in English and French, despite the fact that a large proportion of such studies are carried out in China and South-East Asia and are therefore (also) published in Asian languages. Finally, the geographical distribution of the reviews in Table 2 clearly posits the possibility of a Eurocentric bias of the biomedical discourse.

### Publication Date

This variable is particularly interesting for the PHA category, as articles cover, uniquely, four decades of research, over which the dominance of pharmacology has radically evolved. The two earliest articles [52,53] published before 2000, stand out as *overviews* and recommend to combine anti-inflammatory drugs (non-steroidal analgesic) with other medicaments, notably painkillers (paracetamol) or immunosuppressors (methotrexate, hydroxychloroquine, sulphasalazine as second line drugs for the control of the progression of the disease) as well as other initiatives such as splintage, intra-articular injections, physiotherapy, hydrotherapy, occupational therapy, aids and appliances, psychological management and correct footwear [52], and a multidisciplinary approach with input from orthopaedic surgeons, occupational therapists, physiotherapists, nurses, orthotics and prosthetics departments and chiropodists [53]. Interestingly, at that point in time the authors point out that despite the hope placed in immunosuppressive and combination treatments there is no clinical evidence to prove that the evolution of the disease has been halted. The articles evoke new biological treatments as a prospect for the future.

The four articles [51,54–56] published between 2000 and 2006 are *systematic reviews* and generally much more detailed in the description of RA as a systemic inflammatory autoimmune disease. Despite the richness and complexity of the text, no words are spent to describe the relationship with other approaches: no other type of treatment is mentioned beyond pharmaceutical therapy that becomes the only reference. Coherently with the hopes evoked in the two earliest articles, the focus is on new therapies and strategies (*sawtooth* [54]), with particular emphasis on the prospects that the new biological drugs appear to offer, especially if started at the early stages of the disease.

The last two articles [57,58] are also *systematic reviews*, and describe the most recent pharmacological approaches in terms of molecules’ combinations and aims of the therapy (avoiding recurrence, improving quality of life and achieving *remission* [58]) as the only viable option. No doubt is raised about the medical legitimacy and, while non-pharmacological therapies are mentioned they are not discussed.

This concise excursus shows how anti-inflammatory drugs articles stand out as the dominant approach: not only, in fact, the available biomedical literature exceeds by far in numerosity other approaches, also, these articles tend to present the therapy as the dominant and so, *conventional* for RA. So much so, that by the years 2000 the very meaning of *remission* appears to be altered in the clinical practice. In fact, while “remission is defined as either the reduction or disappearance of the signs and symptoms of a disease” (Wikipedia: remission (medicine)), the very low rate of actual remission (20-25% [59]), moved the acceptable end points of the therapy to “low grade disease activity”, creating a fluid area where RA *remission* is used to indicate the stabilization of the disease, i.e. the halting of its progression [56], “remission-like state” [59], not its reduction below the threshold of activity (DAS28 ≤2.2).

The dominant position of PHA emerges also in the other articles of our selection that frame PHA more or less explicitly as the reference. The pharmacological approach is presented often in the opening section as the treatment which it seems necessary to deal with. Also, with few exceptions (acupuncture [37] and massage [60]) clinical trials present pharmacological control cohorts, despite highlighting limitations and (severe) adverse effects, which are often the argument to present the existence a multiplicity of therapeutic approaches [61,62].

Other articles do not so much aim at presenting the impact of a single complementary therapy, but tend to oversee a plurality of approaches [43,62]. In such cases they even go as far as discussing the impossibility of an actual *remission*.

The year of publication gives also another insight for these therapies, as some of them seem to be outdated for RA, e.g. antibiotics, electrostimulation and laser show very early publications with no follow-up, mirroring (for antibiotics) discontinued research for unsuccess or possibly indicating, lack of interest and/or of fundings.

## Automatic learning – systemic approach

Although the above analysis focuses on one variable at a time, it almost always highlight recurrences or influences from other variables. In order to extract additional relationships that may not emerge directly from manual curation, machine learning approaches are applied for a systemic analysis of Table 2 from which Date, Geography and Background have been removed (see Methods and Supporting Information S1 Fig).

With this reduced representation we attempt to address two major questions: (i) how socio-anthropological variables can help framing the biomedical categories (i.e. what keeps biomedical categories distinct in a socio-anthropological frame, see Section *supervised analysis*), and, complementary to this, (ii) how socio anthropological variables can help identifying common ground among biomedical categories (i.e. what makes biomedical categories similar in a socio-anthropological perspective, see Section *unsupervised analysis*).

### Supervised analysis – socio-anthropological biomedical categories

We were interested in assessing whether there exists a common (or *reference*) socio-anthropological representation of each biomedical category. An intuitive approach to solve this issue is given by a simple *majority voting* algorithm, i.e. given all the articles belonging to one biomedical category, we identify its representative vector as the one having for each of the six variables the value that represents the absolute majority (i.e. the three rows of Fig 3, see Methods). As shown in Fig 3, visual column-wise exploration enables to easily grasp unique and shared characteristics of each category.

**Figure 3.**
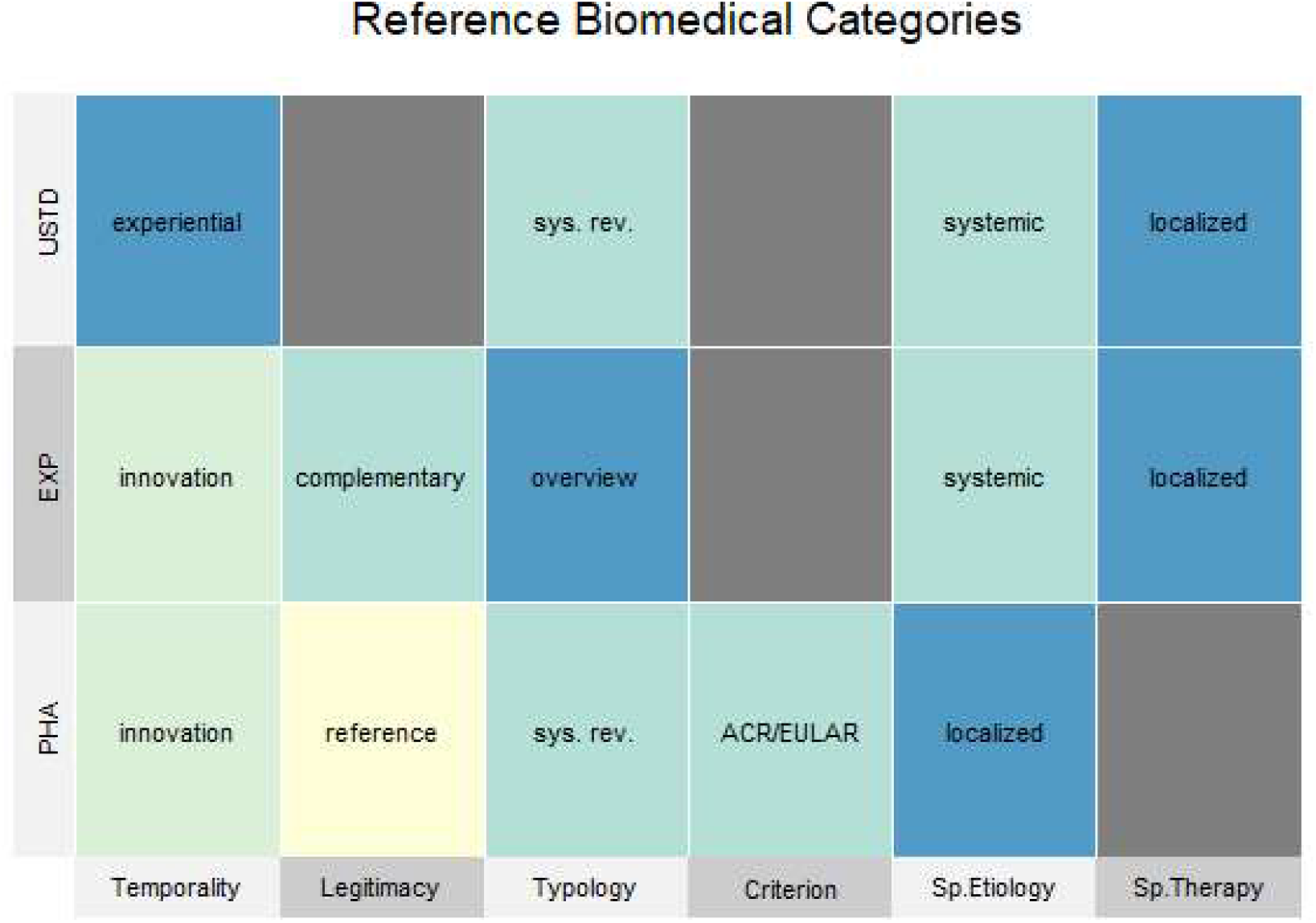
Vectorial representation of the medical discourse referred to the three biomedical UnSTanDardized, EXPerimental and PHArmacological categories of approaches using socio-anthropological categories. Each row of the above table represents a biomedical category in the form a six-dimensional vector, where each dimension contains the representative socio anthropological value for the corresponding category. The representative value is obtained as the majority frequency, across all representations within a category. Colors are only used to distinguish elements, with no other meaning, their values is directly labeled in the cells of the table. Cells in gray represents NA values, i.e. variables for which there is no majority, in other words there is no dominant value for this variable.

**PHA** is uniquely *Legitimized* as the *reference*, dominated by the *ACR/EULAR Criterion* and by a *localized spatialization of the etiology*, while the conceptualization on the *spatialization of the therapy* is neglected or left ambiguous (NA). While the two former results are expected given the role of the ACR/EULAR in biomedicine, the latter may be somewhat surprising given the emphasis that state-of-the-art medicines (Preventive Predictive Personalized and Participatory Precision, hence 3-5P medicines [11]) give to the systemic approach (from systems to network medicine [13,14]) able to prevent diseases via early diagnoses with observation of variables that go beyond localized symptoms [63,64].

**EXP** is the only category dominated by the *overview Typology* of articles. This is expected, and understandably due to the definition of *experimental*, which implies that there exist only a limited number of the prior studies needed to compile systematic overviews. This category is also majorly defined as *complementary* (Legitimacy). This is quite interesting, as, despite EXP approaches being both *innovative* and (partially) *systemic*, two features that are more in line with the ambitions of state-of-the-art/future medical approaches as mentioned above, this, globally, does not move EXP in the medical discourse from an ancillary therapeutic position of *complementary* approaches to PHA.

**USTD** is uniquely characterized by the lack of a dominant diagnostic criterion and by an *experiential Temporality*. The former is interesting as USTDs do not exclude standards, but simply seem to freely choose among available options, disregarding the dominance of ACR/EULAR. The latter reflects two aspects that boost each other: on the one side the lack of recent USTD reviews (see S1 Table) makes it difficult to assess whether more recent reviews might have touched upon the latest findings in basic biology (the inflammatory reflex, or the relation between mechanotransduction and inflammation [65–67]), and at the same time highlight the discontinued scientific production of reviews on USTD. Whether this is due to the lack of fundings offered by biomedicine dominated by PHA, or to an inertia from within the USTDs to modify the perception of Temporality cannot be assessed easily, but might be worth further exploration.

Shared values are also relevant, and interesting is the fact, also, that our approach enables a direct comparison between EXP and USTD, that never cross reference nor mention each other in the body of the articles.

**EXP-PHA** share *innovation*, thus possibly representing the future of medicine, with the former complementing the latter.

**USTD-PHA** share the *Typology* dominated by *systematic reviews*, that is however representative of two very different situations, better explained by the analysis done on Background: in PHA the authorship is dominantly of rheumatologists. This likely contributes to the unity of views presented in this category, and, also, well identifies the closed circle of experts, clearly overlapping with the rheumatology specialty. RA however is defined as an autoimmune musculoskeletal disorder, possible integration of these expertise may still fit within the biomedical paradigm, while opening to a broader conception of the malady. USTD authorship being dominated by non-rheumatologists and mostly relating about techniques they do not apply themselves, directly affects the scientific narrative and its conclusions.

**EXP-USTD** share the conception of the spatialization of the disease characterized by a *systemic etiology* and a *localized therapy*, somewhat highlighting the hiatus between the theorization of the progression of medicine and its clinical practice, where systemic conceptualizations are more easily found in therapeutic approaches that are outside of the dominant paradigm (PHA). While the date of publication must be taken into account to correct for this trend, we can observe that the recent articles of our selection (from 2017 on) still confirm this observation.

### Unsupervised analysis - emergent properties from the medical discourse

Given the overall small samples (28) and space (6) size, we chose the scatter plot represented in Fig 4 to position the articles in a 6-dimensional space and let emerge visual clusters that represent groups of articles sharing coherent socio-anthropological coding, beyond the boundaries of their biomedical categorization. In the following a description of the groups observed in Fig 4 is offered quadrant by quadrant, counterclockwise.

**Figure 4.**
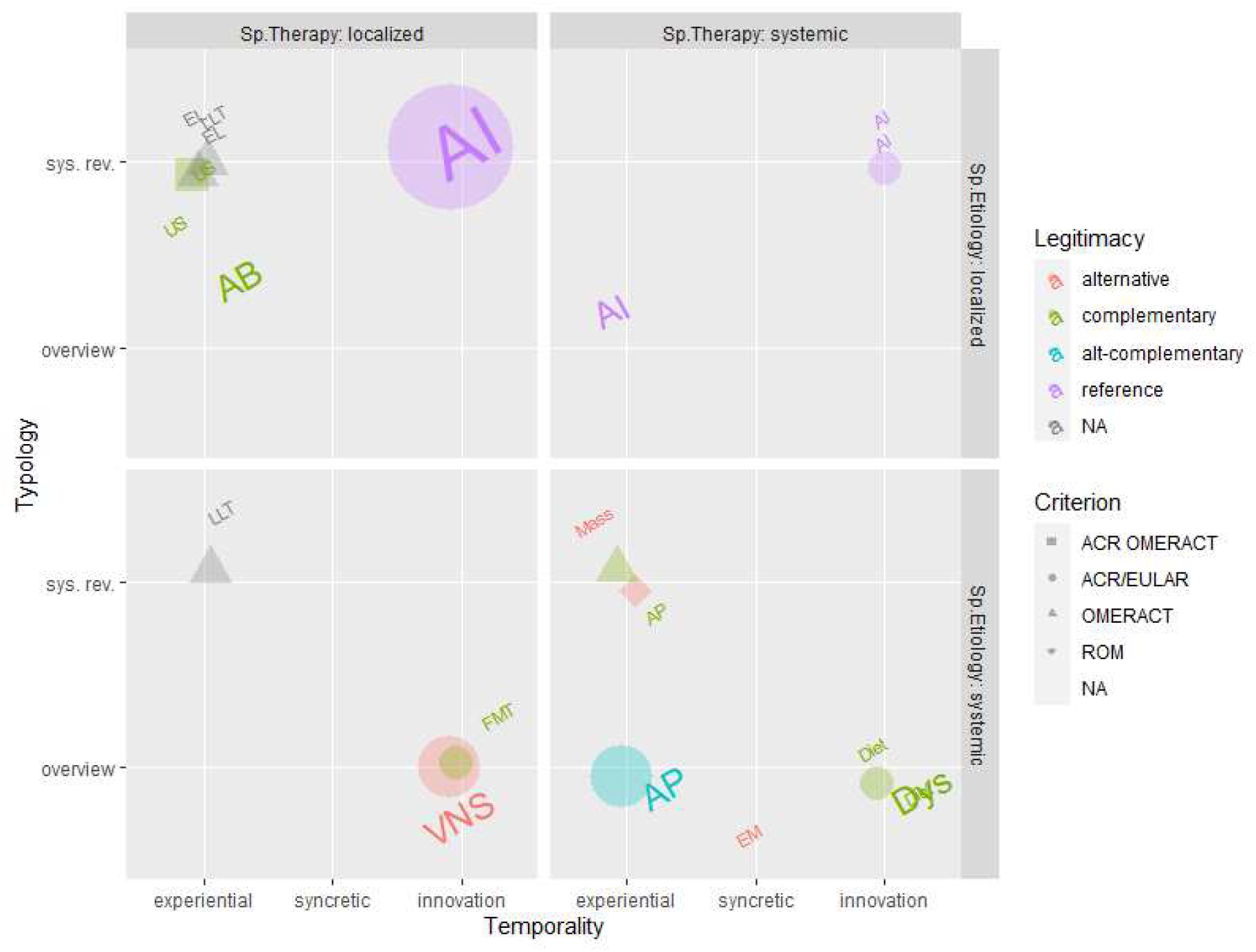
Graphical representation of 6 of the variables under study. Each point whose size is proportional to the number of occurrences in our articles selection, and whose color refers to the Criterion adopted represents a therapy characterized. Labels on the points are taken from Table 2. Starting from the top right side (Quadrant I) counterclockwise to the bottom right side (Quadrant IV) several groups (clusters) are described, representing emergent properties of the medical discourse.

**Quadrant I**, recollects therapies that share a *localized etiology* and a *systemic therapy* conception, unique to PHA. In the earliest reviews of our selection ([52–55] see Table 2) RA is indeed considered a localized disease, i.e. focusing on joints and cartilage, whose treatment however is considered to be systemic.

**Quadrant II** contains therapies characterized by a coherent *local* spatialization. This spatialization is characteristic of the most recent PHA [51,56–58], which evolves its perception of the etiology of the disease from systemic (Quadrant I) to localized, and by the group of *complementary* USTDs (US, EL, LLT). *Localized etiology* for PHA means translating the pathological mechanism in the molecular interactions between molecules, cells and tissues, whereas for USTD it means locating the disease where the body hurts.

Quadrant II is clearly broken (left from right) by Temporality as well as Criterion (green gray versus purple) and Legitimacy (all shapes versus circles). While the right hand side includes only Anti-Inflammatory (Ais), on the left hand side stand therapies that root their strength on *experience* and freely refer to a variety of diagnostic or no standards (OMERACT, ACR/EULAR in combination or none, all shapes of green/gray dots). These characteristics are also accompanied by being ancillary (*complementary* Legitimacy) approaches to PHA. This category includes not only USTD, as already observed in the supervised analysis, but also the obsolete antibiotic approach (AB).

In particular the clustering approach enables to observe that USTDs refer in this Quadrant solely to the use electrical and optical stimuli, completely distinct from mechanical and electromagnetic stimuli, all in quadrant IV, i.e. with an opposite spatialization. EXP therapies never fall in Quadrant II, that is to say, as mentioned already, that the most innovative therapies are not localized.

**Quadrant III** is poorly populated: systemic etiology and localized therapy are rare medical representations. They include however the cutting edge approaches of VNS and FMT, the only EXP legitimating themselves as *alternative*. This offers another viewpoint on the barrier that separate categories, already appearing with the analysis of the Background (rheumatology vs others), indicating in opposing spatializations (local vs systemic) an additional category for segregation.

Finally, **Quadrant IV**, densely populated, can also be subdivided into two clusters: one collecting all mechanically (and EM) stimulated USTDs, and one collecting *complementary* EXP. Here PHA never appears, i.e. it is never associated to a coherently systemic representation of the malady. The same observation done in Quadrant II returns, regarding the distinction of USTDs by localization (II and IV quadrants) with types of stimuli. While magneto- [68,69], mechano- [70,71], electro- [72,73] and opto- [74,75] sensing are recognized abilities of all our cells, mechanotransduction appears to be framed exclusively in a coherently as a localized spatialization, and presents itself as *alternative*, while electrical and optical stimuli require a coherently systemic spatialization, with diversified *legitimacy*.

## Discussion

From the point of view of the textual analysis, strong heterogeneity of the analyzed literature was observed, regarding contents, form and authors. This heterogeneity carries along hierarchical relationships. They concern not only the rather predictable centrality of the gold standard treatment (PHA), but also a whole series of mechanisms that generate asymmetries. In general, approaches closer to biomedicine are depicted as more promising than experiential unstandardized approach. When analyzed from a socio-anthropological perspective, a clear dichotomy appears between articles written by teams directly involved in research into a particular therapy or more generally interested in a therapy and articles written by authors who are not specialists in the field. The interest in the background of the authors therefore showed that it was possible to apply the anthropological categories of *etic*, meaning a point of view *external* to the culture studied (here the therapy reviewed), from an *emic* point of view, internal to the culture under study [76]. Interestingly, an **article using an *emic* or *etic* perspective can affect the way a treatment is framed, and overall overlaps with the concept of *overview* (emic) and *systematic review* (etic)**. This distinction allowed us to observe form another point of view the articles (*systematic reviews*) concerning numerous USTD, in most cases edited by the Cochrane Musculoskeletal group, whose members’ background is only in one case explicitly affiliated to a rheumatology department. These articles are statistical meta-analyses that appear to be more objective being based exclusively on clinical trials, the standard approach to produce evidence in contemporary medicine. Yet, subtle biases can be traced in these reports. In fact, although randomized, double blind placebo controlled clinical trials are the most statistically challenging and are hence considered the gold standard [43], the Harmonized Tripartite Guide [77] (which standardizes worldwide the statistical approaches behind clinical trials) offers a plurality of study designs. This is necessary to accommodate not only non-pharmacological treatments, that, for example, barely deal with blinding, but also degenerative conditions, for which placebo controls are not ethical (non-inferiority clinical trials are in this case viable alternative model design). Yet, this statistical categorization is sometimes erroneously translated into a quality ranking and/or associated to lack of standardization, which represents a different and independent issue. As a consequence, we observe that meta-analyses are run discarding what are in fact appropriately designed non-inferiority trials, biasing the meta-analysis conclusion as it is the case when comparing the articles on acupuncture [34–36]. In the case of acupuncture, moreover, two out of three reviews show a clear Eurocentric bias in the choice of selecting only articles written in English, despite the fact that this technique originated and is much more widespread in Asia.

With respect to the automatic learning analysis interesting patterns have emerged. *Spatialization*, for instance, seems a useful concept to navigate the relationships existing among the biomedical categories.

As it currently emerges from our analysis, USTDs share a coherent spatialization (be it *local* or *systemic*), suggesting that these approaches refer to a unified representation of the mechanism of disease and healing. This can be explicitly found in some medical practices that define the medical action as a mean to remove the obstacles that provoke the disease (consider energy stagnation in Traditional Chinese Medicine), a different conceptualization from PHA, which operates downstream of processes gone awry (along the inflammatory cascade for instance), with less emphasis on the causes of the disease. Interestingly, this observation also emerges in the results of anthropological research on CAM. Several studies show that in the discourse of conventional physicians practicing unconventional medicine (CAM) the global or holistic approach (which chooses to deal with the causes of the pathologies more than with the symptoms) is one of the central reasons of their choice of therapeutic approach [78,79].

Currently, state-of-the-art conceptualizations of biomedicine (3-5P medicines [11,80]) focus on the importance of systemic approaches, which implies a *systemic* spatialization of both the *etiology*, going beyond the genetic and integrating environmental factors (and their molecular surrogates), and the *therapy* with a particular emphasis on lifestyle and nutrition. Nevertheless, in our analysis, a coherent *systemic spatialization* is promoted by ancillary (complementary) nutritional therapies (EXP) and mechano-based non-pharmacological approaches (USTD), while state-of-the-art therapies (recent AI and *alternative* EXP) promote a localized vision of the therapy. Further, given the *legitimacy* they refer to (*reference* or *alternative*), as it currently stands one approach seems to attempt to replace the other, with the same type of spatialization. It is therefore a possible first operable recommendation from this work that **transition to** a systemic/holistic approaches promoted by **future 3-5P medicines are likely to require <u>explicit</u> attention, discussion and revisitation of the concepts that frame the perceived spatialization of the etiology of a disease**.

It is also worth focusing on the distinct spatialization that USTD mechanosensing (and magnetosensing) imply versus electric and optic signals. Mechanosensing and mechanotransduction have long been known to impact tissues beyond the single cell, given the continuity with the extracellular matrix [81], naturally implying the necessity for a systemic approach. This is ignored in the AP reviews but mentioned in the review on massage [47], thus explaining the apparent surprising *legitimacy* of this article (*alternative*). Possibly, the growing availability of 3D-bioprinting may clarify the relevance of the environment and hence of the systemic approach also for stimuli of different nature [82], thus offering the scientific bases for translation into clinical practice. Overall however, **the basic biological knowledge on the transduction on physical stimuli fails to be taken into account in USTDs**, a fact that **require a change in the paradigm that approaches therapeutic stimuli**. This may occur by explicit health policy decisions, or implicitly driven by technology. Just like biology and medicine renovated themselves triggered by the technological progress of high-throughput biology [13], slowly integrating informatics to design the current network medicine paradigm [15], integration of physics and engineering dragged by 3D bioprinting may give birth to a more complete biophysical medicine.

Strictly replicating to these observations are the ones descending from *Temporality*. EXP and USTD are represented very differently. The former are optimistically described as promising techniques that are based on a wealth of findings from current research and turned to the future. In contrast, the latter are systematically described as being based solely on experience with very limited attempts to integrate biological research and experiential practices. Acupuncture provides a very clear illustration of this situation: while meta-analyses are generally characterized by purely statistical considerations, with no reference to the therapeutic nor biological rationale, we observe in [36] an attempt to forcefully refer to the biological mechanisms underlying the therapy using “*electroatropines”* propagating a concept presented once [83] in the ‘80s with no other relation to modern experimental biology.

This consideration calls for a two-way revisitation of the perception of USTD: from outside and from within. In fact, while clearly the example above illustrates a bias towards the reliability of USTDs, it is intriguing to observe from a manual search (which did not emerge from our automated queries) that a similar and opposite approach can be found in more recent literature. As it may have appeared already VNS and electroacupuncture share a very clear ground: electrical stimulation of the vagus nerve. It is therefore interesting to observe that the most recent (2018-2021) reviews on RA and AP consist of : a meta-meta-analysis (data are extracted from prior meta-analyses, [84]) conducted in South-America, reporting the overall lack of impact of such therapy on RA, and two meta-analyses conducted in Asia [85,86] describing, conversely, positive impact of the treatment. Only [85] was an accessible open source and to the best of our knowledge rigorously designed study. Still, the limited space for discussion given to the biological mechanism include two research articles published on national or specialized literature (Acupuncture Research [87,88]) disregarding the knowledge on the inflammatory reflex and on other associated biological findings [65,89]. This is even more intriguing as a number of scholars of Chinese origin promote on high-impact journals in basic research the connection between the most recent findings in neurophysiology and acupuncture [90].

Clearly, in this case, differently from the recommendation to integrate multiple Backgrounds to enhance integration from a Spatialization viewpoint, Geography, or better the medical background, may be the focus of integration to overcome the Temporality dichotomy, for example a **multicentric east-west study of acupuncture may lead to innovative conclusions**.

*Legitimacy* offers yet another standpoint: while we defined *alternative* as a relative concept and were able to identify therapies presenting themselves as potentially self-contained, this is not the case for *complementary*, as therapies are always complementary to the reference PHA, and never to the other two classes. To date therefore *complementary* therapies are in fact ancillary to PHA in a clear hierarchical relation. There is therefore room to explore balanced complementarity between EXP, USTD and PHA, likely playing on timing, i.e. exploiting (the prevalence of) different categories at different stages of the disease (prevention, early/acute stages, chronicity). This prioritization may benefit from being guided by cost-effectiveness, exploring the non-pharmacological modalities of USTD and EXP delivery to maximize safety, dissemination and ease of use. This approach may open to a third suggestion, i.e. the development of **the novel area of *therapy* (versus drug) *repurposing***, by **exploiting the numerous existing biomedically approved devices born to deliver USTDs in novel clinical studies** which design should be **guided by the biological mechanisms at the roots of physical transduction and EXP therapies**, in potentially extremely cost-effective ways. Only embryonic is indeed the interaction elicited by VNS and electroacupuncture, knowing for instance that auricular acupuncture stimulates a branch of the vagus nerve in the only anatomical location where it emerges on the surface of the body [91]. A recent publication offers a case for overcoming this compartmentalization [92]. Finally, not foreign to these issue is the distribution of the diagnostic criteria adopted across biomedical categories, possible measures to take into account this observation include, but are not limited to, expanding the variables that contribute to the calculation of DAS28, including qualitative variables mirroring systemic aspects of well-being, like for instance sleep quality, mood, ability to heal wound [63,64,93].

*Typology* also gives room for considerations, meta-analyses should not be constrained by Background nor Geography, and in particular it may be relevant to integrate VNS and TENS or AP studies given the right conditions (stimulation of points in proximity of the vagus nerve), and more control should be done to ensure that the **full range of approved studies design** is appropriately included in **meta-analyses**, a somewhat trivial, but not unnecessary recommendation.

Overall, we observe that the medical discourse is a pluralistic space able to accommodate perspectives that are very heterogeneous. At the same time, the relationships among the different approaches are hierarchical, being the unstandardized therapies marginalized and the pharmacological treatment the dominant one, and this can result in a lack of dialogue, making all this explicit may be the first step towards a novel integration.

## Conclusions

First of all, the limitations of this work are to be highlighted: as mentioned already both the sample size and the space of investigation can be expanded or modified. This is not only true with respect to the categories we focused on, but also with respect to the analyses we chose for this pioneering exploration. Different types of supervised analyses, and even different choices of mapping between the axes and the socio-anthropological variables could let emerge different clusters. We observe however that several of the core pattern emerge across all our (limited number of) analyses letting suppose that they may be the strong enough to be considered for further investigation.

Second, how to use this type of analysis, and if and how they are meaningful and practical cannot be decided at this point. Certainly, medical specialties compartmentalization has a clear reflection on the scientific discourse, impinging on conclusions owing to input data bias. While this aspect is more obvious, the observation that overcoming this obstacle may for example require a reflection on the spatial perception of the disease is way less evident.

Our suggestions are only starting points, but we hope that their dissemination within circles of state-of-the-art medicine (EPMA www.epmanet.eu ; the H2020 and HE ICPerMed initiatives www.icpermed.eu/, the Network Medicine Global Alliance www.network-medicine.org, to name a few) may trigger other richer analyses, whose results may provide tangible support to the offer of cost-effective means to protect citizens by the ever growing burden of NCDs.

## Data Availability

All data produced in the present work are contained in the manuscript and accompanying supplementary material

