## Supplementary Material for "Semi-automated socio-anthropologic analysis of the *medical discourse* on rheumatoid arthritis: potential impact in public health"

### Supplementary Information

Christine Nardini[1*],Lucia Candelise[2,3*], Mauro Turrini[4*], Olga Addimanda[5]

[1] Consiglio Nazionale delle Ricerche, Istituto per le Applicazioni del Calcolo "Mauro Picone", Roma, IT
[2] ISS, Istitut Sciences Sociales, Université de Lausanne, CH
[3] CEPED, Centre Population et Développement, Université de Paris, FR
[4]Institute of Public Goods and Policies (IPP), Spanish National Research Council (CSIC), Madrid - ES
[5]UOC Medicina Interna ad Indirizzo Reumatologico, Ospedale Maggiore, AUSL Bologna, IT

***Key-words:*** non-communicable diseases; non-pharmacological therapies; chronic inflammation; rheumatoid arthritis; health policy; medical socio-anthropology; machine learning; Sustainable Development Goals.

**Literature additional filtering criteria**

The queries on Pubmed were run on February 4th 2020. Before analyzing the articles selected by Pubmed MeSH search, additional filtering has been applied on titles alone in order to refine the selection. The first filtering is a semi-automatic step, the second is manual.

Semi-automatic filtering to remove the following items (color coded in Supplementary Table 1):

- Non RA: reviews not on RA
- Non english: articles in other languages
- Non appropriate: different topic
- Too specific: RA with specific comorbidity; case reports; non systemic approach to RA (i.e. specific joints only)
- Too general: relevant for RA also, but not specific to RA (green)
- Included: the article already falls in another category, we kept it in the most appropriate one
- Misclassified: belongs to another category, where it is not present (orange)
- Older version: when two versions are available we keep the most recent

Second manual curation:

- Identification title-wise of the most general articles, coherent with the subsection of interest. This, in particular for the most abundant category of drug therapy anti-inflammatory, leads to the arguable exclusion (*too specific*) of any article focusing on any broad subset of drugs (DMARDs, chemotherapics etc.)
- Identification where possible of time series (mostly *drug therapy anti-inflammatory; dietary supplement; anti-microbial*), the former for example traces back the evolution of therapy from the introduction of biologics onwards, and the latter morphs into the more general *gastrointestinal microbiome* upon discovery of its relevance in autoimmunity.
- Filtering by availability (availability of the online article in general, and open access afterwards)
- Cochrane reviews are preferred when offered by PubMed ranking by all above criteria.
- Control on specific bias: There exist ample controversy against the usage of sham acupuncture as a mean to include blinding in random control trials (RCT) for acupuncture [1,2]. Opponent to this forceful blinding claim that all points on the body surface are to some extent sensitive to the needle stimulation and therefore comparison

is meaningless/inappropriate. Beyond the controversy, there exist internationally standardized [3] statistical design to model situations where placebo is not possible (this, incidentally, includes degenerative diseases like RA is): *non-inferiority clinical trial*s, that compare the gold standard (pharmacological therapy in this case) with the same therapy plus an additional approach, acupuncture in this case. Given that the two automatically selected papers [4] and [5] include sham acupuncture and/or exclude non-inferiority trials, to offer a more complete analysis of the medical discourse we added the article by Wang et al. [6] published on the same year as [5] and analyzing the same data (7 out of 8 total trials are the same), including active control trials and analyzing them separately from RCT.

**Data representation**

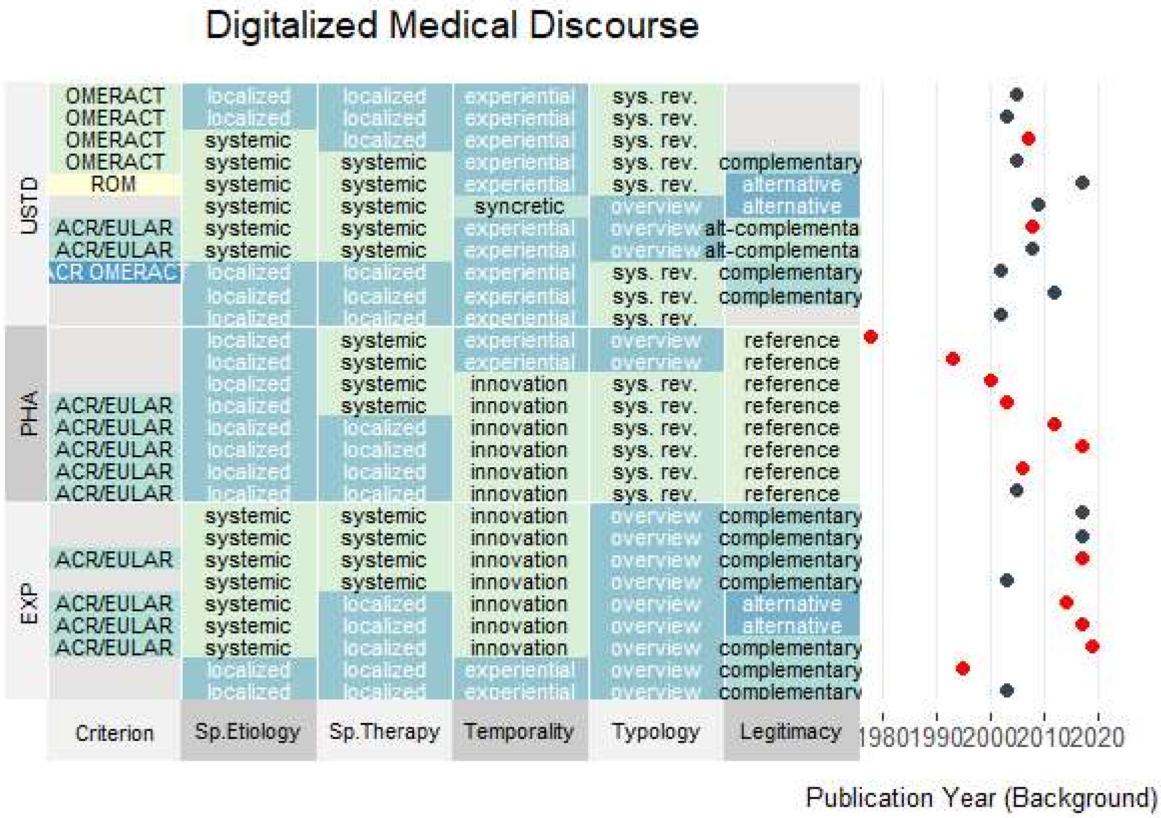


**Figure S1** *Representation of the digitalized medical discourse in a heatmap, emphasizing socio-anthropological variables, versus biomedical categories. Colors are only used to distinguish elements, with no other meaning, their values is directly labeled in the cells of the table. Light gray values are NA. Adjacent plot represents the corresponding Publication Year. Dots in red represent main authors' Background in rheumatology, in black everything else. Geography is omitted given the relatively uninformative content (Eurocentric). The*

*majority of Rheumatologists in PHA versus the two other categories is visually clear, as is the extent of the temporal articles distribution.*
